# Integration and scale-up of a primary healthcare-based chronic wound care package for persons affected by skin-NTDs and other conditions in Ethiopia: a protocol for an implementation research study

**DOI:** 10.1101/2024.06.12.24308591

**Authors:** M Kinfe, M Semrau, A Mengiste, O Ali, T Ajeme, SA Bremner, N Hounsome, V Anagnostopoulou, M Brewster, L Rugema, Eiman Siddig Ahmed, Agumasie Semahegn, A Fekadu, G Davey

## Abstract

**Introduction:** The profound impact of wounds on the quality of life of those affected is often underestimated. Chronic wounds impose substantial burdens on individuals and communities in terms of disability, mental distress, stigma, and economic productivity losses. To effectively address these challenges, an integrated and comprehensive approach to primary healthcare-based chronic wound care prevention and management is essential. This implementation research study aims to assess the integration and scale-up of a comprehensive package of primary healthcare-based wound care and psycho-social support for persons affected by chronic wounds caused by neglected tropical diseases (NTDs) and other conditions in selected districts in Ethiopia.

**Methods:** The study will be implemented in Central Ethiopia in three stages, utilizing a mixed-methods approach to co-develop a comprehensive care package and progressively implement the care package building on learnings from successive stages of implementation. Stage 1 will encompass the co-development of a holistic wound care package and strategies for its integration into routine health services. Stage 2 will involve a pilot study in one sub-district, to establish the care package adoption, feasibility, acceptability, fidelity, potential effectiveness, readiness for scale-up, and costs. Stage 3 will involve the scale-up of the wound care package and its evaluation in several districts.

**Ethics and dissemination:** Ethics approval was obtained for the study from the relevant authorities in both the UK (Ref no: ER-BSMS9D79-6) and Ethiopia [Reference no. 013/23/CDT]. The results of the study will be disseminated through a variety of channels, including publications in scientific journals, conference presentations, policy briefs, and workshops. This will ensure that the findings are disseminated widely to the scientific community, policymakers, and the public.

**Strengths and limitations of this study:** *Strengths:*

- This study will address a critical gap in Ethiopia, where there is a significant health burden due to chronic wounds from skin-NTDs and other conditions, for which primary healthcare integration of wound care could improve accessibility and outcomes for affected persons, their families and communities.
- The study will use implementation research methods to examine the integration and scale-up of a primary healthcare-based package for wound care and psychosocial support, which is crucial for wider adoption and sustainability.
- The use of a mixed-methods design will enable a comprehensive understanding of the care package’s feasibility and effectiveness.

*Limitations:*

- Lack of a comparison group (i.e. control group).
- Whilst the study will include provision of essential medical supplies to bridge local shortages and support disadvantaged patients, provision of care after the study period is beyond the scope of this study. However, the study team will work closely with the local health administration and the Ministry of Health to ensure the sustainability of services.

## INTRODUCTION

Many Neglected Tropical Diseases (NTDs) can cause wounds and other skin complications. Wounds are among the most frequently encountered skin problems in rural settings in low- and middle-income countries (LMICs), where skin NTDs are also endemic (1, 2). Wounds are among the major causes of visits to hospitals in Africa, accounting for 30–42% of hospital attendance and 9% of deaths every year (3). However, they are also among the most underreported health challenges in many parts of Africa, probably partly because of limited access to health care services. Over 60% of the population of the African continent resides in rural areas, with up to 80% facing geographical barriers to accessing modern healthcare (4) .

The causes of wounds are numerous, varying markedly by age group, environment, and occupation. The causes range from trauma, burns and bacterial infections to non-communicable diseases (NCDs) such as peripheral arterial disease and diabetes (1, 2). Fifteen to twenty five percent of patients with diabetes develop a diabetic foot ulcer in their lifetime (5, 6) and up to one-third of patients in acute and long-term care develop a pressure ulcer (7). Wounds in most parts of Africa are caused by NTDs, assaults, road crashes, occupation-related injuries (e.g., at construction sites, on farms, and domestic accidents), animal bites, burns, surgery, and diseases resulting in acute or chronic soft tissue damage. Well-known examples of NTDs that can cause wounds and other skin complications in Ethiopia are leprosy, cutaneous leishmaniasis, lymphatic filariasis (LF), podoconiosis, onchocerciasis, snakebite, and guinea worm, as well as ectoparasites like scabies and tungiasis.

Studies investigating wounds in resource-limited settings are scarce. While chronic wounds impose huge burdens on affected individuals and communities in terms of disability (14), mental distress (15), depression (10), stigma (16–19), and loss of economic productivity (14, 20–22)a wound can have on the quality of life of those affected is therefore under-recognized (8–13). Furthermore, findings from some cost analyses carried out in high-income countries (HICs) show a strikingly high cost burden (23, 24), and this also needs to be assessed in resource-limited settings.

High-quality and timely wound management is vital to prevent infections and disabilities. Unfortunately, wound management is often expensive and ineffective in the areas where NTDs occur. In these settings, the number of health professionals is limited, health workers are minimally trained in providing curative care, and self-care is not sufficiently implemented. Skills in evaluating abnormal signs and symptoms, such as when to stop using topical antiseptics, when to suspect secondary infection and when to suspect malignant alteration are lacking. There is limited training and the need for expertise in wound care is not well recognized. A study conducted in Ghana and Benin interviewing healthcare personnel dealing with the wound management of Buruli ulcer patients reported that standards of wound care differed greatly both between personnel and between institutions (25). Variations in wound care may derive from a lack of formal wound care education, limited high-quality evidence, inconsistent guidelines, and the diversity of clinical techniques used to support wound healing.

Multiple gaps in care have also been identified in Ethiopia, including limited physician knowledge of wound care, and poor use of best practices (26). A survey conducted among nurses in Ethiopia found their knowledge and practice of wound care to be poor. The study found important gaps in knowledge and alarming lack of evidence-based practice. In addition, unfavourable attitudes among nurses towards wound care were identified. Updating nurses’ knowledge and creating favourable attitudes through training on prevention and management of foot care was recommended (6). This might be achieved through provision of structured pre-service and in-service training and education programs on wound care. Establishing and putting in place wound healing and care guidelines and assessment tools in health facilities and conducting further qualitative research on the levels of wound care practices is also recommended (26).

Integrated wound management is focused on sustainable prevention and care of wounds in settings with limited resources. It comprises identification, treatment and preventive measures. Wound care is a cross-cutting component in the management of a number of skin-NTDs (such as leprosy, Buruli ulcer, yaws, cutaneous leishmaniasis, tropical ulcers, LF and podoconiosis) and can be delivered through the same individuals in the same health care settings. In many co-endemic areas, leprosy and Buruli ulcers are managed in the same facilities (27). In Ethiopia, an integrated approach for LF and podoconiosis morbidity management has been demonstrated to be feasible (28).

Consultation with affected persons and implementer groups in Ethiopia suggests that development and evaluation of an integrated wound management service applicable across skin conditions would be enormously beneficial not just for persons affected by podoconiosis, LF and leprosy, but also for those with cutaneous leishmaniasis, diabetes and injuries. This is in line with the cross-disease emphasis of the new World Health Organization (WHO) Roadmap for NTDs (29).

Innovative ways of ensuring support, including use of mHealth tools, tele-dermatology, and online training courses, may also be relevant. These innovative technological methods can help to bridge the gap between the burden of skin diseases (including chronic wounds) and the lack of capable staff in resource-poor settings by bringing essential health services from central settings closer to peripheral levels (30). Opportunities for low-cost tele-dermatology applications will continue to increase, making it feasible for any provider in any setting to be connected to a tertiary hospital dermatologist, surgeon or other appropriate specialist. Incorporating telehealth into wound care delivery not only facilitates more timely patient access, but can be a more efficient utilization of wound specialists (31). Recent literature indicates that telehealth facilitates earlier assessment and treatment of wounds and improves patient outcomes. Telehealth may be especially beneficial to people who reside in rural areas (32, 33) because it may reduce their need to travel long distances (and thus decrease travel-related costs) to specialty wound care clinics (34). The use of telehealth for wound care may also reduce the overall number of clinic visits that patients require, which, in turn, may reduce related costs to the health care system (35) .

A comprehensive review of published papers in the field of chronic wound care revealed a low proportion of global publications with high quality, robust information (36). Our observation identifies a number of issues relating to the management of chronic wounds in Ethiopia including:

- The continued use of outdated practices which did not promote healing;
- Limited access for patients to effective treatment that would heal their wound;
- Lack of a coordinated approach to chronic wound management.

Shortcomings of existing services include a lack of evidence-based practice, insufficient resources, and in particular, poor understanding of the true cost to society and service providers.

In Ethiopia, high priority needs to be given to persons affected by chronic wounds. This is therefore an opportune time to address the challenge of integrating a comprehensive package of chronic wound care interventions into routine health services. The Health Extension Programme, rolled out successfully in Ethiopia since 2006, now boasts more than 38,000 community-based Health Extension Workers (HEWs) and a supervisory system to support them. Their reach has been extended through the women-centred Health Development Army (HDA), members of which link ’model families’ with five other households to implement health initiatives (37). These cadres are ideally placed to offer simple, low-tech wound care and psychosocial support to people living with diseases associated with chronic wounds in Ethiopia (38), and to refer patients who need more specialised wound care. The Ethiopian Ministry of Health (MoH) is aware of the nature and scale of the problem and has requested implementation research to guide two ‘axes’ of integration: i) development of an integrated package of physical and psychosocial care interventions for persons with chronic wounds, and ii) subsequent integration of this comprehensive wound care package into government-run health services.

To address this, the ‘Excellence in Disability Prevention Integrated across NTDs’ Part II (EnDPoINT II) implementation research study is running in Ethiopia from 2021 to 2025, funded by the National Institute for Health and Care Research (NIHR) in the UK.

The aim of this implementation research study is to integrate and scale up a comprehensive package of primary health care-based physical and psychosocial care for persons affected by chronic wounds due to NTDs and other conditions in selected districts in Ethiopia across three project stages.

## METHODS AND ANALYSIS

### Conceptual framework for study

This study is an implementation research study in that: its research questions have a strong focus on implementation strategy; it includes a wide range of implementation outcomes; and it is being conducted in real-world settings and populations (39). The conceptual framework used for the study will be the Medical Research Council (MRC) framework for complex interventions (40), embedded within a ‘Theory of Change’ approach . The MRC framework for complex interventions has been used widely and proposes four phases in the development and evaluation of interventions:

i. development,
ii. feasibility/piloting,
iii. evaluation, and
iv. implementation

These phases are considered to be an iterative rather than a linear process. ‘Theory of Change’ has been defined as a “theory of how and why an initiative works” (41); it enhances the MRC framework for complex interventions through its theory-driven approach, which can be incorporated into and provide practical guidance for the different phases of the MRC framework (41). Within this study, the ‘Theory of Change’ approach will only be used within two of the four phases of the MRC framework: the development and piloting phases; for these two phases, there is good evidence for the practical feasibility and usefulness of an embedded ‘Theory of Change’ approach (42), whereas this is not yet the case for the phases on implementation and evaluation.

### Study design

Stage 1 corresponds to Phase 1 of the MRC framework on complex interventions – the development of the comprehensive wound care package. Various activities will be carried out to inform the development of the care package and strategies for its integration into the routine health care delivery system. A scoping review (of peer-reviewed literature) and grey literature review will be conducted to gain an understanding of publications relevant to the development of the comprehensive wound care package. A situational analysis and resource assessment will be conducted to provide the local context for the development and implementation of the care package in the selected districts in Ethiopia, and to identify the resources available for the implementation of the care package. Three ‘Theory of Change’ workshops will be held with members of the EnDPoINT II research team and key stakeholders, to identify the desired outcomes, indicators, interventions, and measurement of the outcomes for the wound care package, and to encourage stakeholder buy-in to the study. Further, key informant interviews and focus groups will be carried out with key stakeholders, to assess the care package’s feasibility (i.e., the extent to which the intervention can be carried out within the routine health system), acceptability (i.e., the perception among stakeholders that the comprehensive wound care package is agreeable), and appropriateness (i.e. the perceived fit or relevance of the care package to key stakeholders). Informed by the above steps, the comprehensive wound care package consisting of physical and psychosocial care interventions at the healthcare administration, health facility and community level will be developed.

Stage 2 corresponds to Phase 2 of the MRC framework on complex interventions, the piloting of the comprehensive wound care package in one selected sub-district in Ethiopia. This will enable study of the factors that influence integration of the care package and assessment of the wound care package’s adoption, feasibility, effectiveness, fidelity, and coverage within the government-run health system and the roles of key health system actors.

Stage 3 corresponds to Phases 3 and 4 of the MRC framework, and will involve scale-up of the wound care package in several districts in Ethiopia based on learnings from the previous two stages.

### Setting

The EnDPoINT II study is planned to be implemented in two zones: East Gurage in Central Ethiopia and Awi in Amhara Region north-western Ethiopia. Based on the 2007 census conducted by the Central Statistical Agency of Ethiopia (CSA), Gurage has a total population of 1,280,483 (44). The six largest ethnic groups reported in Gurage Zone were the Gurage people (82%), the Mareqo or Libido (4.3%), the Amhara (3.4%), the Kebena (3.3%), the Silt’e people (2.7%), and the Oromo (1.7%); all other ethnic groups made up 2.6% of the population. Gurage was spoken as a first language by 80.5% of the population, 5.3% spoke Amharic, 4.1% spoke Libido, 3.2% spoke Kebena, 3% spoke Silt’e, and 1.1% spoke Oromo; the remaining 2.8% spoke other primary languages. The majority of inhabitants (51%) were reported to be Muslim, while 41.9% practised Ethiopian Orthodox Christianity, 5.8% were Protestants, and 1.3% Catholic. Seventy-nine percent (79%) of all eligible children were enrolled in primary school, and 12% in secondary schools (44).

Awi is one of the zones in the Amhara Region of Ethiopia. Based on the 2007 CSA, this zone has a total population of 982,942. The two largest ethnic groups reported in Agew Awi were the Awi (59.8%) a subgroup of the Agaw, and the Amhara (38.5%); all other ethnic groups made up 1.7% of the population. Amharic was spoken as a first language by 53.4% of the population, and 45% spoke Agew-Awigna; the remaining 1.6% spoke other primary languages. 94.4% practiced Ethiopian Orthodox Christianity, and 4.5% of the population said they were Muslim. Seventy-two percent (72%) of all eligible children were enrolled in primary school, and 16% in secondary schools (44).

### Sample Selection

For Stage 1, as no primary research will be conducted for the literature reviews and situational analysis, no participants will need to be recruited for these aspects (though some of the information for the situational analysis/resource assessment may be obtained by contacting key officials, if necessary, for example by phone). Members of the EnDPoINT II research team in Ethiopia participated in both ‘Theory of Change’ workshops held to date. Participants of the second ‘Theory of Change’ workshop included members of the wider EnDPoINT II Consortium, and other key stakeholders such as current non-communicable disease (NCD) and NTD experts (including NGO implementers), policy-makers, health service planners, health workers, community leaders, and members of affected persons’ organisations. Participants in the key informant interviews and focus groups during Stage 1 will include key stakeholders who already are, or who will be involved as part of the implementation of the study, in the provision and/or receipt of NCD/NTD care, for example dermatologists, surgeons, HEWs, NGOs working within NCDs and NTDs, primary health care workers, health managers and decision-makers, policy-makers, community leaders, traditional and religious leaders, and affected persons and their families.

During the pilot study and scale-up of the care package in Stages 2 and 3, a wide range of stakeholders at each of the three levels of the health system (health care organization, health facilities, and the community) will receive training to implement the various components of the care package. This will include staff at the health care organization level, zonal and district level health office staff, psychiatric nurses, senior health care workers, senior pharmacists, pharmacy staff, health centre staff (health officers and nurses), HEWs, and members of the community (see ‘Procedure’ section below for further details). They will also be invited to participate in pre-post knowledge, attitudes and practices (KAP) questionnaires, and an evaluation of the delivery of the training.

In addition, affected members of the community, i.e. people with chronic wounds, as well as their families and their communities will be included in the study as recipients of the interventions. Affected people may also be involved in some of the delivery of the training packages or interventions as experts by experience, in order to provide a ‘lived testimony’ of living with chronic wounds and include a social contact element in the training and interventions. In addition, both affected persons (patients) and community members will take part in a quantitative pre-post survey, which will be administered to each of these two groups before and after implementation of the care package (with various follow-up points). Furthermore, to gain deeper insight into the lived experiences of persons affected by chronic wounds as well as their caregivers, in-depth qualitative interviews will be conducted with them alongside the quantitative data collection.

Adults aged 18 or over will be included in the study. All aspects of the study will be conducted in either Amharic or English, depending on which language is most appropriate for each participant group. When necessary, all study materials, including participant information sheets and consent forms, will be translated into Amharic, the official language of Ethiopia, which is spoken and understood by the majority of people in the country, including those in the study areas.

### Sampling and sample size

Key stakeholders will be identified and recruited using purposive sampling techniques for the ‘Theory of Change’ workshops in Stage 1, the key informant interviews and focus groups in Stages 1 and 2, and the training elements in Stage 2. Participants will be selected based on their role and position. The EnDPoINT II research team will initially contact key stakeholders to invite them to participate. Snowballing techniques may also be used, where each identified key stakeholder will be asked to recommend other potential participants. The number of participants taking part in the ‘Theory of Change’ workshops in Stage 1, and the key informant interviews and focus groups during Stages 1 and 2 of the study, will be guided by the number of key stakeholders that are identified. A sample size calculation is not appropriate for these qualitative data collection methods but we will rather follow a data saturation approach, i.e. data collection will continue until sufficient information has been obtained or where further interviews fail to generate new themes. However, the number of participants during the ‘Theory of Change’ workshops and focus groups will not exceed 12 participants at a maximum, to ensure that all participants have the opportunity to speak. For the training elements in Stage 2 of the study, the number of participants will be established based on the number of people who are suitable and available for training.

For the before-and-after (pre-post) collection of quantitative data in Stage 2 of the study through the patient and community surveys (with data collection points at baseline before implementation of the care package, and at 3- and 12-months follow-up), paired data from 120 patients will be collected in the selected sub-districts; 60 from each district (45). An allowance for 5% attrition will be made, implying that 126 patients will need to be sampled pre-training. For the community survey, 200 participants will be included. These numbers will provide good precision of estimated retention proportions that will be used to inform the sample size and other design elements in Stage 3 of the study (45).

### Inclusion/exclusion criteria

Participation in each of the sub-studies will be voluntary. Participants in the second ‘Theory of Change’ workshop, the key informant interviews and focus groups, those who are taking part in the training evaluations, and participants of the patient and community surveys, will be required to give their informed consent to take part (at baseline). If a person does not consent to take part in any of these research evaluation activities, that person will be excluded. Participants will all be over 18 years of age. Participants must have adequate hearing to ensure effective communication. Individuals with severe hearing impairments that significantly hinder communication will be excluded. Participants must be in a stable health condition, meaning they are not acutely ill or experiencing severe pain that could interfere with their ability to participate fully in the study. Individuals who are currently experiencing acute illness or severe pain will be excluded.

#### Intervention package description

The interventions included in the care package will be organised according to the three levels of the health care system in Ethiopia: the health care organization level which includes the Ministry of Health, Regional Health Bureau, zonal health department and district health office; the health care facility level which includes primary hospitals, health centers and health posts; and the community level. The draft care plan may include the following interventions:

| Domains | Descriptions |
| --- | --- |
| Clinical photography in chronic wound care using smartphones | There is a high level of smartphone ownership and usage among nurses and health officers working at health centres and primary hospitals. These health workers will take clinical photographs after obtaining written consent from the patient mainly to gain advice from peers/consultants and for treatment and disease monitoring purposes. They will photograph the wound on initial assessment and repeat every four weeks or more frequently if the wound condition changes rapidly. The consulting hospital wound care practitioner will ask the HC nurse practitioner a series of basic wound assessment questions. A treatment plan will be developed based on visual examination of the wound and responses to the wound assessment questions |
| Health care organization level: | • High-level awareness-raising and mobilization: participatory workshop for zonal and district health bureau officials and community advisory board;<br>• Programme management, i.e. working in partnership with key health care organization staff to implement the necessary structures and budget for the delivery of the care package;<br>• Capacity-building, i.e. ‘Training of Trainers’ (ToT) for health care organization staff about wound care provision and supervision. Resources from the WHO (46) and other sources (38) provide rich information about managing wounds;<br>• Formation of Public Advisory Group with members of the community, NTD and NCD task force. |
| Health care facility level: | • Awareness-raising amongst health facility staff and attendees: i) workshop for facility-based staff by NTD/NCD focal person at health care organization level; ii) posters in health centres; iii) health education sessions by health facility staff for attendees of health facilities;<br>• Case detection, assessment, and treatment and wound care initiation by health officers and nurses, including supply package, psychosocial support, and health education for patient self-care;<br>• Capacity-building for facility-based health centre staff;<br>• Supervision and mentoring for health centre staff;<br>• Use of telehealth digital technologies, mobile health apps, and remote patient monitoring to support long-distance consultation on clinical health care;<br>• Institution-based rehabilitation, e.g. reconstructive surgery, links with multi-disciplinary team, internist, surgeons and mental health care providers;<br>• Supply chain management. |
| Community level: | ○ Community awareness-raising: awareness-raising workshops for members of the community by HEWs and NGOs;<br>○ Information dissemination in the community by HEWs and NGOs (information leaflets and posters in health posts; posters/loudspeaker in the marketplace) on the common causes of chronic wounds;<br>○ Active case detection by HEWs;<br>○ Continuing care/follow-up home visits to patients and their families by HEWs;<br>○ Community-based rehabilitation, including home visits, workshops, patient associations and self-help groups;<br>○ Capacity-building for HEWs;<br>○ Supportive supervision for HEWs. |

### Procedures

#### Stage 1

##### Literature reviews

Three reviews will be conducted. The first will be a scoping review on the implementation of primary health care and community-based physical and psychosocial interventions for the management of chronic wounds and lessons learnt from program implementation in low- and middle-income countries. Secondly, a systematic review will be carried out on telehealth in regard to barriers and challenges to telemedicine/health for chronic wound care in LMICs. Finally, a grey literature review will be conducted to support the development of the wound care package and strategies for its integration into the routine health care delivery system in Ethiopia. The wound care package will therefore draw on publications listed in bibliographic databases (through the scoping and systematic reviews), as well as grey literature (through the grey literature review), and include:

- existing wound care training packages and materials from both LMICs and HICs;
- documents relevant to the primary care provision of physical or psychosocial care for wounds, such as relevant project reports, programme documents and relevant publications in scientific journals;
- any other relevant materials.

##### Situational analysis and resource assessment

To understand better the context of implementation and resources that may be available to support the implementation of the interventions, standard situational assessment and resource mapping tools will be used. The tools will be developed based on expert input from members of the EnDPoINT II Consortium and from previous situational analysis and mapping tools that have been used in other projects such as EnDPoINT I (47).

##### ‘Theory of Change’ workshops

The main aim of the workshops is to build consensus and define the desired outcomes, indicators, interventions, assumptions and measurements for the comprehensive chronic wound care package, and to encourage stakeholder buy-in to the study (41). The map will be developed through a series of workshops (at least three) with all key stakeholders that will include policy makers, implementers, care providers, service users, community leaders and organizations. An indicator matrix will be developed, describing the type of indicator (input, process, output or outcome), the method of assessment and the source of data.

The ‘Theory of Change’ map developed during the workshops may need to be adapted further following the key informant interviews and focus groups, to account for the information provided on the wound care package’s feasibility, acceptability and appropriateness. Similarly, the ‘Theory of Change’ developed during Stage 1 may need to be adapted following pilot-testing of the care package during Stage 2.

##### Key informant interviews and focus groups

The main aim of the key informant interviews and focus groups conducted in Stage 1 of the study will be to assess the feasibility, acceptability and appropriateness of the draft wound care package, including the key assumptions identified during the ‘Theory of Change’ workshops (41).

#### Stage 2

##### Pre-piloting in a health facility

An initial pre-pilot will be carried out during Stage 2, to assess the main training elements of the health facility and community components of the draft wound care training package, and evaluate their feasibility, adoption, acceptability, fidelity, effectiveness, coverage, their readiness for piloting, and the suitability of the training materials.

Training of health officers and nurses will include delivery of integrated wound care and psychosocial support; training of people with chronic wounds in self-care; awareness-raising for health centre attendees; detection and assessment, treatment and care initiation; referral to providers of institutional-based rehabilitation; and enhancement of institutional culture around continued learning. Training materials will include Chronic Wound Management Guidelines, the Skin-NTDs app (WHO/NLR), and simple wound management and self-care using online training materials, dermatological packages, guides and tele-dermatology (46).

For the HEWs, the training will include: delivery of wound care for people with chronic wounds, including training patients in self-care; referrals to health facilities; community awareness-raising and mobilization; active case detection; continuing care; community-based rehabilitation; and monitoring and evaluation.

Training materials at the community level will include pictorial pocket booklets, posters based on type of skin lesions/wounds, pictorial pocket booklets and posters promoting self-care. Tasks for HEWs will include screening community members for suspected skin-NTDs and other diseases with chronic wounds, community education, and advice on self-care and prevention of recurrence for people affected by chronic wounds (46).

The training materials will be evaluated and revised through change in knowledge, attitudes and practices (KAP) questionnaires, and an evaluation of the delivery of the training.

The topic guides and interview questions for the key informant interviews and focus groups, and the questionnaires for evaluation of the training, will be developed once Stage 1 has been completed and the materials for the training will be developed based on the ToC, the narrative literature review, and the systematic/scoping reviews.

The draft care package will be revised further based on the results of the pre-piloting before being piloted in one sub-district.

##### Pilot in sub-district

All wound care package interventions will be piloted at all three levels of the health system in one sub-district. As a first step, this will involve training of health professionals and other stakeholders within the district at each level of the health system, including:

- ‘Training of Trainers’ (ToT) for staff at the health care organization level, to enable them to deliver training to zonal and district level health office staff in wound care for people affected by chronic wounds due to NTDs and other conditions. The ToT will be delivered by a Master trainer.
- ToT for staff at the health care organization level, to enable them to deliver training to zonal and district level health office staff to supervise facility-based health officers, nurses and HEWs. The ToT will be delivered by a Master trainer.
- Zonal and district level health office staff, to enable them to train and supervise facility-based health officers, nurses and HEWs. This training will be facilitated by staff at the health care organization level (i.e. recipients of the above ToTs), and will involve training in supervision, as well as wound care for people with NTDs and other conditions.
- Health centre staff (i.e. health officers and nurses), trained by zonal and district level health office staff.
- A multi-disciplinary team comprising senior health care workers on chronic wound management, and senior pharmacists, to enable them to provide clinical mentoring for facility-based health officers and nurses. This training will involve training in mentoring, as well as wound care for people with chronic wounds due to NTDs and other conditions.
- Pharmacy staff who manage and receive wound care supplies in health facilities. The training will cover supply chain management and will be facilitated by a pharmacist.

All training elements (in comprehensive chronic wound care, supportive supervision and supply chain management) will be evaluated through pre-post change in KAP questionnaires, and an evaluation of the delivery of the training. The questionnaires for evaluation will be the same ones used during pre-piloting, though these may be refined and improved based on the pre-piloting findings. Based on the evaluations, the training materials may be improved further ahead of the scale-up in Stage 3.

Following training, all interventions included in the wound care package (as outlined in the ‘Key interventions within the care package’ section above, refined further based on the Stage 1 activities) will be implemented, to assess their adoption, feasibility, potential effectiveness, fidelity, and coverage. This will be assessed qualitatively through observation, through key informant interviews and focus groups with stakeholders who received the training and delivered the interventions, and with recipients of the interventions. We will also collect before-and-after quantitative data on the number of cases identified and treated by health facility staff and HEWs, and patient- and community-level outcomes. To provide a preliminary estimate for contact coverage in the pilot district, the number of cases identified and treated will be divided by the total population in need of services (this latter data will be taken from previous prevalence studies).

#### Stage 3

After assessing the implementation scalability and sustainability, Stage 3 will be conducted. This stage corresponds to Phases 3 and 4 of the MRC framework on complex interventions, i.e. the evaluation and implementation. We will scale up the wound care package and its evaluation in two selected districts in Ethiopia. Scale-up will be informed by the results of Stages 1 and 2 of the study.

##### Economic Analysis

The health economics analysis aims to i) Estimate resources and costs associated with development, piloting and integrating the new wound care package into the routine health care system, and ii) Assess the cost-effectiveness of this care package using a range of clinical and quality-of-life outcomes.

In Stage 1 (the development of the wound care package) we will collect data on resources involved in the development of the wound care package, i.e. situational analysis, **‘**Theory of Change’ workshops, key informant interviews, focus groups and preparation of training materials. The costs will cover staff time spent on developing guidelines and training materials, and expenses associated with conducting interviews and workshops (e.g. venue hire, travel expenses, accommodation, food, digital equipment and stationary materials). Data will be collected using purpose-designed questionnaires and interviews with programme managers.

In Stage 2 (piloting of the wound care package) data will be collected on implementing the care package in one sub-district in Ethiopia. This will include training of health officers and nurses in delivery of integrated wound care and psychosocial support, detection and assessment, treatment and care initiation, referral to providers of institutional-based rehabilitation, training of people with chronic wounds in self-care, awareness-raising for health centre attendees and enhancement of institutional culture around continued learning. We will collect data on the costs of training (e.g. trainers and trainees time, training materials, digital equipment, travel and accommodation); leaflet and poster preparation and distribution; wound treatment supplies (e.g. soap, ointment, bandages, gloves, gauzes, antiseptics and antibiotics); storage and delivery of treatment supplies, and telehealth gadgets. Data will be collected from financial documentation, purpose-designed questionnaires and interviews with programme managers.

In Stage 3 (scale-up of the wound care package in two selected districts in Ethiopia) we will collect data to inform the cost-effectiveness analysis of the implementation of the wound care package. The data will include resources and costs related to the delivery of the intervention and the use of healthcare services used by people with chronic wounds. The services will include contacts with healthcare specialists and traditional healers, hospitalisations, medication, traditional remedies, and out-of-pocket expenses related to the management of chronic wounds. We will collect data on the number of days when individuals were completely unable to work because of their wounds (or go to school) and the number of days when they experienced some difficulties when working (or attending school). We will also collect data on money borrowed by families over the past year to cover costs of treatments and living. A cost-effectiveness analysis will be performed using a range of clinical and quality-of-life outcomes collected during the implementation research study to inform decision-makers whether the new care package provides good value for money.

##### Data management and analysis

All data analyses will be conducted by members of the EnDPoINT II research team at BSMS and CDT-Africa, AAU. For data collection and storage, we will use REDCap, a secure web application for building and managing online surveys and databases. Data quality control processes will be agreed and implemented.

The documents identified through the literature reviews will be summarised systematically into a narrative review and will be used to guide the development of the wound care package. Data from the situational analysis and resource assessment will be described quantitatively as frequencies or qualitatively (free-text data). A ‘Theory of Change’ map and an accompanying narrative will be produced based on the ‘Theory of Change’ workshops.

Key informant interviews and focus group discussions will be audio-taped, transcribed in Amharic and then translated into English with independent translation checks of selected interviews. The data will be analysed using thematic analysis with the assistance of a qualitative software package (NVivo). Validity of data will be assessed through the identification of initial codes by an external experienced qualitative researcher, agreement over the framework through which the data will be presented, presentation of initial findings to experienced academics to assess plausibility, and single counting of identified events. Validity assurance will also involve checking of alignment of research questions, interview guides and sampling procedures. The observation data collected during Stage 2 will be recorded on paper and will be analysed in a similar way. During Stages 2 and 3, quantitative data will be collected on the REDCap secure web platform.

Participant progress through the pre-post study will be shown using a flow diagram according to the CONSORT Statement 2010 extension for pilot and feasibility studies (48) adapted to reflect the non-randomised nature of our study. Data will be summarised descriptively using means and standard deviations for normally distributed variables, medians and interquartile ranges for skewed continuous variables and frequencies and percentages in each category for categorical variables. Amount of missing data will be summarised; no imputation will be conducted. Change from baseline in patient-centred outcomes and other point estimates will be presented with 95% confidence intervals. Success criteria will be agreed at appropriate point in the project and will be evaluated at each stage and appropriate protocol modifications will be made as necessary. A full statistical analysis plan will be agreed prior to database lock and final analysis.

Wound care clinics will be set up in health facilities in two selected districts in Ethiopia. Setting up this new service will involve the review of all wound assessment tools, patient documentation, referral forms and patient management systems and audit processes. A plan for evaluating the service will be developed using data (e.g. healing rates, wound size improvement, origin of referrals, number of infections and client satisfaction). The results from each of the activities in Stages 1 and 2 will be triangulated to develop and finalise the wound care package and strategies for its integration into the routine health care delivery system in selected districts/zones in Ethiopia.

## Data Availability

All data produced in this study will be available upon reasonable request to the authors.

http://www.cdt-africa.org

## ETHICS AND DISSEMINATION

### Ethics

Ethical approval for the study has been obtained from the Brighton and Sussex Medical School Research Governance and Ethics Committee (BSMS RGEC) in the UK (Stage 1 ref: ER/BSMS9D79/6; Stage 2 ref: ER/BSMS9D79/7) and the Institutional Review Board of the College of Health Sciences at Addis Ababa University in Ethiopia (013/23/CDT). All participants, who must be over 18 years of age, will be provided with a participant information sheet. Those who choose not to participate will be excluded from the study. Participants retain the right to withdraw from the study at any time until data have been aggregated without providing a reason or facing any negative consequences, including those related to their care, treatment, or employment.

Maintaining the confidentiality and anonymity of participant data is paramount throughout the study, encompassing data collection, storage, analysis, and publication. Multi-layered safeguards will be meticulously implemented to protect the sensitive nature of the information collected and preserve the anonymity of participants. Personal identifiers will be utilized instead of names, and identifying data will be securely stored separately from the corresponding codes used on the data collection sheets and databases. Only pseudonymised data will be subjected to data analyses. All data will be securely stored in a restricted OneDrive folder, accessible solely to authorized members of the research team.

### Dissemination

A publication plan has been developed outlining the dissemination of study findings through various channels. The plan encompasses the publication of multiple peer-reviewed articles in scientific journals. Additionally, outcomes will be shared with a broader audience, including the scientific community, policymakers, healthcare professionals, and study participants. Dissemination will likely take the form of an end-of-study workshop, the distribution of a summary document, and the development of policy briefs highlighting key findings.

## Authors’ contributions

This manuscript was a collaborative effort by the EnDPoINT II research team, with all authors contributing to its development and approving the final version. MK led the writing of the manuscript. MS and AM led the development of the study protocol. GD and AF are the Principal Investigators of the EnDPoINT II study. OA, AM, MK, MS, SAB, MB, TA, AS, LS,ES, VA and NH are part of the core EnDPoINT II research team. OA, AM, TA and MK are responsible for the implementation of the study in Ethiopia. SAB is responsible for overseeing the statistical elements of the study, NH leads the economic aspects, and VA is the study statistician.

## References

1. Toutous Trellu L, Nkemenang P, Comte E, Ehounou G, Atangana P, Mboua DJ, et al. Differential Diagnosis of Skin Ulcers in a Mycobacterium ulcerans Endemic Area: Data from a Prospective Study in Cameroon. PLoS neglected tropical diseases. 2016;10(4):e0004385.

2. Ryan T. Public health dermatology: Regeneration and repair of the skin in the developed transitional and developing world. International journal of dermatology. 2006;45:1233–7.

3. Norman R, Matzopoulos R, Groenewald P, Bradshaw D. The high burden of injuries in South Africa. Bulletin of the World Health Organization. 2007;85(9):695–702.

4. Amzat J, Razum O. Rural Health in Africa. In: Amzat J, Razum O, editors. Towards a Sociology of Health Discourse in Africa. Cham: Springer International Publishing; 2018. p. 109–24.

5. Rice JB, Desai U, Cummings AK, Birnbaum HG, Skornicki M, Parsons NB. Burden of diabetic foot ulcers for medicare and private insurers. Diabetes care. 2014;37(3):651–8.

6. Abate TW, Enyew A, Gebrie F, Bayuh H. Nurses’ knowledge and attitude towards diabetes foot care in Bahir Dar, North West Ethiopia. Heliyon. 2020;6(11):e05552.

7. Reddy M, Gill SS, Rochon PA. Preventing pressure ulcers: a systematic review. Jama. 2006;296(8):974–84.

8. Effah A, Ersser SJ, Hemingway A. Support needs of people living with Mycobacterium ulcerans (Buruli ulcer) disease in a Ghana rural community: a grounded theory study. Int J Dermatol. 2017;56(12):1432–7.

9. Narahari SR, Bose KS, Aggithaya MG, Swamy GK, Ryan TJ, Unnikrishnan B, et al. Community level morbidity control of lymphoedema using self care and integrative treatment in two lymphatic filariasis endemic districts of South India: a non randomized interventional study. Transactions of the Royal Society of Tropical Medicine and Hygiene. 2013;107(9):566–77.

10. Bartlett J, Deribe K, Tamiru A, Amberbir T, Medhin G, Malik M, et al. Depression and disability in people with podoconiosis: a comparative cross-sectional study in rural Northern Ethiopia. International health. 2016;8(2):124–31.

11. Bakhiet SM, Fahal AH, Musa AM, Mohamed ESW, Omer RF, Ahmed ES, et al. A holistic approach to the mycetoma management. PLoS neglected tropical diseases. 2018;12(5):e0006391.

12. Wiese S, Elson L, Feldmeier H. Tungiasis-related life quality impairment in children living in rural Kenya. PLoS neglected tropical diseases. 2018;12(1):e0005939.

13. Walker SL, Lebas E, De Sario V, Deyasso Z, Doni SN, Marks M, et al. The prevalence and association with health-related quality of life of tungiasis and scabies in schoolchildren in southern Ethiopia. PLoS neglected tropical diseases. 2017;11(8):e0005808.

14. Bekri W, Gebre S, Mengiste A, Saunderson PR, Zewge S. Delay in presentation and start of treatment in leprosy patients: a case-control study of disabled and non-disabled patients in three different settings in Ethiopia. International journal of leprosy and other mycobacterial diseases : official organ of the International Leprosy Association. 1998;66(1):1–9.

15. Mousley E, Deribe K, Tamiru A, Tomczyk S, Hanlon C, Davey G. Mental distress and podoconiosis in Northern Ethiopia: a comparative cross-sectional study. International health. 2015;7(1):16–25.

16. Hofstraat K, van Brakel WH. Social stigma towards neglected tropical diseases: a systematic review. International health. 2016;8 Suppl 1:i53–70.

17. Tora A, Davey G, Tadele G. A qualitative study on stigma and coping strategies of patients with podoconiosis in Wolaita zone, Southern Ethiopia. International health. 2011;3(3):176–81.

18. Tora A, Franklin H, Deribe K, Reda AA, Davey G. Extent of podoconiosis-related stigma in Wolaita Zone, Southern Ethiopia: a cross-sectional study. SpringerPlus. 2014;3:647.

19. Person B, Bartholomew LK, Gyapong M, Addiss DG, van den Borne B. Health-related stigma among women with lymphatic filariasis from the Dominican Republic and Ghana. Social science & medicine (1982). 2009;68(1):30–8.

20. Tekola F, Mariam DH, Davey G. Economic costs of endemic non-filarial elephantiasis in Wolaita Zone, Ethiopia. Tropical medicine & international health : TM & IH. 2006;11(7):1136–44.

21. Perera M, Whitehead M, Molyneux D, Weerasooriya M, Gunatilleke G. Neglected patients with a neglected disease? A qualitative study of lymphatic filariasis. PLoS neglected tropical diseases. 2007;1(2):e128.

22. Martindale S, Mkwanda SZ, Smith E, Molyneux D, Stanton MC, Kelly-Hope LA. Quantifying the physical and socio-economic burden of filarial lymphoedema in Chikwawa District, Malawi. Transactions of the Royal Society of Tropical Medicine and Hygiene. 2014;108(12):759–67.

23. Guest JF, Ayoub N, McIlwraith T, Uchegbu I, Gerrish A, Weidlich D, et al. Health economic burden that different wound types impose on the UK’s National Health Service. International wound journal. 2017;14(2):322–30.

24. Tchero H, Kangambega P, Lin L, Mukisi-Mukaza M, Brunet-Houdard S, Briatte C, et al. Cost of diabetic foot in France, Spain, Italy, Germany and United Kingdom: A systematic review. Annales d’endocrinologie. 2018;79(2):67–74.

25. Velding K, Klis SA, Abass KM, Tuah W, Stienstra Y, van der Werf T. Wound care in Buruli ulcer disease in Ghana and Benin. The American journal of tropical medicine and hygiene. 2014;91(2):313–8.

26. desta m, Tenaw M, Ayalew E. Level of Knowledge and Wound Care Practice at a Tertiary Referral Hospital in Ethiopia: A Survey in 180 Nurses. 2020.

27. Walsh DS, De Jong BC, Meyers WM, Portaels F. Leprosy and Buruli ulcer: similarities suggest combining control and prevention of disability strategies in countries endemic for both diseases. Leprosy review. 2015;86(1):1–5.

28. Deribe K, Kebede B, Tamiru M, Mengistu B, Kebede F, Martindale S, et al. Integrated morbidity management for lymphatic filariasis and podoconiosis, Ethiopia. Bulletin of the World Health Organization. 2017;95(9):652–6.

29. WHO. Ending the neglect to attain the Sustainable Development Goals A road map for neglected tropical diseases 2021–2030.

30. Mieras LF, Taal AT, Post EB, Ndeve AGZ, van Hees CLM. The Development of a Mobile Application to Support Peripheral Health Workers to Diagnose and Treat People with Skin Diseases in Resource-Poor Settings. Tropical medicine and infectious disease. 2018;3(3).

31. Moffatt JJ, Eley DS. The reported benefits of telehealth for rural Australians. Australian health review : a publication of the Australian Hospital Association. 2010;34(3):276–81.

32. Jia H, Cowper DC, Tang Y, Litt E, Wilson L. Postacute stroke rehabilitation utilization: are there differences between rural-urban patients and taxonomies? The Journal of rural health : official journal of the American Rural Health Association and the National Rural Health Care Association. 2012;28(3):242–7.

33. Skolarus TA, Chan S, Shelton JB, Antonio AL, Sales AE, Malin JL, et al. Quality of prostate cancer care among rural men in the Veterans Health Administration. Cancer. 2013;119(20):3629–35.

34. Buzza C, Ono SS, Turvey C, Wittrock S, Noble M, Reddy G, et al. Distance is relative: unpacking a principal barrier in rural healthcare. Journal of general internal medicine. 2011;26 Suppl 2(Suppl 2):648–54.

35. Chanussot-Deprez C, Contreras-Ruiz J. Telemedicine in wound care. International wound journal. 2008;5(5):651–4.

36. Zenilman J, Valle MF, Malas MB, Maruthur N, Qazi U, Suh Y, et al. AHRQ Comparative Effectiveness Reviews. Chronic Venous Ulcers: A Comparative Effectiveness Review of Treatment Modalities. Rockville (MD): Agency for Healthcare Research and Quality (US); 2013.

37. Ayode D, McBride CM, de Heer H, Watanabe E, Gebreyesus T, Tadele G, et al. The association of beliefs about heredity with preventive and interpersonal behaviors in communities affected by podoconiosis in rural Ethiopia. The American journal of tropical medicine and hygiene. 2012;87(4):623–30.

38. Keast DH. World Alliance for Wound and Lymphedema Care. 2020.

39. David H. Peters NTT, Taghreed Adam. Implementation Research in Health: A Practical Guide. 2013.

40. Craig P, Dieppe P, Macintyre S, Michie S, Nazareth I, Petticrew M. Developing and evaluating complex interventions: the new Medical Research Council guidance. BMJ (Clinical research ed). 2008;337:a1655.

41. De Silva MJ, Breuer E, Lee L, Asher L, Chowdhary N, Lund C, et al. Theory of Change: a theory-driven approach to enhance the Medical Research Council’s framework for complex interventions. Trials. 2014;15:267.

42. Breuer E, De Silva MJ, Fekadu A, Luitel NP, Murhar V, Nakku J, et al. Using workshops to develop theories of change in five low and middle income countries: lessons from the programme for improving mental health care (PRIME). International journal of mental health systems. 2014;8:15.

43. De Silva M, Breuer E, Lee L, Asher L, Chowdhary N, Lund C, et al. Theory of Change: a theory-driven approach to enhance the Medical Research Council’s framework for complex interventions. Trials. 2014;15:267.

44. Commission(OPCC) OotPC. The 2007 Population and Housing Census of Ethiopia.

45. Teare MD, Dimairo M, Shephard N, Hayman A, Whitehead A, Walters SJ. Sample size requirements to estimate key design parameters from external pilot randomised controlled trials: a simulation study. Trials. 2014;15:264.

46. Keast DH. Wound and Lymphoedema Management 2nd Edition Focus on Resource-limited Settings. 2020.

47. Semrau M, Ali O, Deribe K, Mengiste A, Tesfaye A, Kinfe M, et al. EnDPoINT: protocol for an implementation research study to integrate a holistic package of physical health, mental health and psychosocial care for podoconiosis, lymphatic filariasis and leprosy into routine health services in Ethiopia. BMJ open. 2020;10(10):e037675.

48. Eldridge SM, Chan CL, Campbell MJ, Bond CM, Hopewell S, Thabane L, et al. CONSORT 2010 statement: extension to randomised pilot and feasibility trials. BMJ (Clinical research ed). 2016;355:i5239.

